# The Impact of COVID-19 on Adjusted Mortality Risk in Care Homes for Older Adults in Wales, United Kingdom: A retrospective population-based cohort study for mortality in 2016-2020

**DOI:** 10.1101/2020.07.03.20145839

**Authors:** Joe Hollinghurst, Jane Lyons, Richard Fry, Ashley Akbari, Mike Gravenor, Alan Watkins, Fiona Verity, Ronan A Lyons

## Abstract

**Background:** Mortality in care homes has had a prominent focus during the COVID-19 outbreak. Multiple and interconnected challenges face the care home sector in the prevention and management of outbreaks of COVID-19, including adequate supply of personal protective equipment, staff shortages, and insufficient or lack of timely COVID-19 testing. Care homes are particularly vulnerable to infectious diseases.

**Aim:** To analyse the mortality of older care home residents in Wales during COVID-19 lockdown and compare this across the population of Wales and the previous 4-years.

**Study Design and Setting:** We used anonymised Electronic Health Records (EHRs) and administrative data from the Secure Anonymised Information Linkage (SAIL) Databank to create a cross-sectional cohort study. We anonymously linked data for Welsh residents to mortality data up to the 14^th^ June 2020.

**Methods:** We calculated survival curves and adjusted Cox proportional hazards models to estimate hazard ratios (HRs) for the risk of mortality. We adjusted hazard ratios for age, gender, social economic status and prior health conditions.

**Results:** Survival curves show an increased proportion of deaths between 23^rd^ March and 14^th^ June 2020 in care homes for older people, with an adjusted HR of 1·72 (1·55, 1·90) compared to 2016. Compared to the general population in 2016-2019, adjusted care home mortality HRs for older adults rose from 2·15 (2·11,2·20) in 2016-2019 to 2·94 (2·81,3·08) in 2020.

**Conclusions:** The survival curves and increased HRs show a significantly increased risk of death in the 2020 study periods.

## INTRODUCTION

### Background

Mortality in care homes has had a prominent worldwide focus during the COVID-19 outbreak [1], [2] but few detailed analyses have been conducted. Care homes are a keystone of adult social care. They provide accommodation and care for those needing substantial help with personal care, but more than that, they are people’s homes [2], [3]. In 2016, there were 11,300 care homes in the UK, with a total of 410,000 residents [4]. Within care homes people live in proximity, and may live with frailty and many different health conditions, making them susceptible to outbreaks of infectious disease [3]. COVID-19 is described by Lithande et al, as ‘…a dynamic, specific and real threat to the health and well-being of older people’ (2020,p.10) [5]. The impacts of COVID-19 on this population group have been reported widely in both international and UK media, and in a growing peer reviewed literature.

Multiple and interconnected challenges face the care home sector in the prevention and management of outbreaks of COVID-19[2]. In the literature, these challenges are reported to include poor access and supplies of personal protective equipment (PPE) for care home staff [1], [2], [6], [7], staff shortages [1], [2], insufficient or lack of timely COVID-19 testing [2],[6] and related clinical challenges as some older adults with COVID-19 may be asymptomatic, or not display expected symptoms [1], [2], [5], [6]. Once there is an outbreak the disease can spread quickly within a care home setting, and be difficult to contain [6]–[8]. A further challenge is in managing the impact of practices to shield care home residents and isolate those who are infected. These practices can result in social isolation from families, friends and communities, with negative impacts on health and wellbeing [2], [5]. Set against these challenges is the caring, innovative, and resilient response of care home staff and residents in managing the situations they face [9].

This confluence of events in the context of the pandemic, and impacts for residents, their families and care home staff, has been framed as a human rights issue [10]. In the UK there is also contestation regarding the implications of underinvestment in the care home sector, and the interface with the health sector, for example, of the rapid hospital discharge policies in the early period of the lockdown [7], [11], [12]. COVID-19 is a rapidly evolving complex issue requiring near real time data, analyses and a multidisciplinary team to devise, implement and evaluate a wide variety of inter- and cross-sectoral interventions to minimise population harm.

The use of existing anonymised routinely collected longitudinal data can help to provide rapid access to large-scale data for studies and provide robust evidence for commissioning decisions and policy [13]. In this study, we utilise the Secure Anonymised Information Linkage (SAIL) Databank [14]–[16] to investigate mortality in care homes in Wales in the initial phase of the UK lockdown, and compare this with corresponding data from the four most recent years to estimate excess mortality.

### Research Questions

We aimed to compare the mortality risk for older care home residents (60+) in Wales for each year between 2016 and 2020. To do this we performed two sets of analyses:

1. How does mortality in care homes for older adults compare between 2016-2020?
2. How does care home mortality for older adults compare between 2020 and 2016-2019 in context of the population of Wales?

## METHODS

### Study design

We used anonymised Electronic Health Records (EHRs) and administrative data from the Secure Anonymised Information Linkage (SAIL) Databank to create a cross-sectional cohort study.

### Data sources

Our cohorts were created using data held within the SAIL Databank [14]–[16]. The SAIL Databank contains longitudinal anonymised administrative and healthcare records for the population of Wales. The anonymisation is performed by a trusted third party, the National Health Service (NHS) Wales Informatics Service (NWIS). The SAIL Databank has a unique individual anonymised person identifier known as an Anonymous Linking Field (ALF) and unique address anonymised identifier known as a Residential Anonymous Linking Field (RALF)[17] that are used to link between data sources at individual and residential levels, respectively. Individual linking fields, nested within residential codes, are contained in the anonymised version of the Welsh Demographic Service Dataset (WDSD), replacing the identifiable names and addresses of people registered with a free-to-use General Practitioner service.

Our cohort of older care home residents was determined by linking to an existing index for anonymised care home addresses from a previous project[18] and utilising the WDSD for address changes. We determined if someone was a care home resident by linking their de-identified address information to the residences indexed as a care home in the WDSD. The anonymised care home index was created using the Care Inspectorate Wales (CIW) [19] data source from 2018 and assigning a Unique Property Reference Number (UPRN) to each address[20]. We included care homes with a classification of either care homes for older adults, or care homes for older adults with nursing in our list. The UPRN was double-encrypted into a project level RALF and uploaded into SAIL to create a deterministic match to the WDSD. From an analysis perspective, both residents and care homes are de-identified prior to any analysis.

### Setting & Participants

To answer our research questions we created separate data sets, both with different settings and participants.

1. Initially we focussed on the phase of the UK lockdown for the COVID-19 pandemic, from March 23^rd^ to June 14^th^ 2020, and compared the mortality risk of care home residents to those in the corresponding period in previous years (2016-2019). Individuals in Wales aged 60+ years identified in the SAIL Databank as a resident in a care home on March 23^rd^ in one of our study years (2016–2020). We created 5 cohorts, one for each year of study, and treated these as independent.
2. We compared the mortality risk of being in a care home at the population level between January 1^st^ 2020 – April 30^th^ 2020 and January 1^st^ 2016 – December 31^st^ 2019. We used the WDSD to create population wide cohorts, this included all individuals resident in Wales from 2016 – 2020. We stratified the dataset in to four sub-groups for comparison:
  a. Non care home residents, resident in Wales on 1/1/2016. Residents were followed up until they moved out of Wales, died, or 31/12/2019.
  b. Non care home residents, resident in Wales from 1/1/2020 to 30/4/2020. Residents were followed up until they moved out of Wales, died, or 30/04/2020.
  c. Care home residents, resident in a care home for older people in Wales on 1/1/2016. Residents were followed up until they moved out of Wales, died, or 31/12/2019.
  d. Care home residents, resident in a care home for older people in Wales from 1/1/2020 to 30/4/2020. Residents were followed up until they moved out of Wales, died, or 30/04/2020.

### Hospital Frailty Risk Score

The Hospital Frailty Risk Score (HFRS) was developed using Hospital Episode Statistics (HES), a database containing details of all admissions, Emergency Department attendances and outpatient appointments at NHS hospitals in England, and validated on over one million older people using hospitals in 2014/15 [21]. The HFRS uses the International Classification of Disease version 10 [22] (ICD-10) codes to search for specific conditions from secondary care. A weight is then applied to the conditions and a cumulative sum is used to determine a frailty status of: Low, Intermediate or High. We additionally included a HFRS score of ‘No score’ for people who had not been admitted to hospital in the look back period. We calculated the HFRS using the Patient Episode Database for Wales (PEDW), the Welsh counterpart to HES, on the entry date for each of our studies, with a two year look back of all hospital admissions recorded in Wales.

### Outcome of interest – Mortality

We used a combination of the Office of National Statistics (ONS) Annual District Death Extract (ADDE), WDSD, and Consolidated Death Data Source (CDDS) to link historic and current mortality information. The CDDS is a combination of the ONS mortality data along with the death records found in the Master Patient Index (MPI), and was used to identify deaths in 2020, the ADDE and WDSD were used to identify deaths from 2016-2019.

### Demographics

Additional demographic information was taken from the WDSD. The WDSD includes the week of birth, the 2011 Lower layer Super Output Area (LSOA) used to assign the 2019 Welsh Index of Multiple Deprivation (WIMD), and gender. We calculated the age of individuals on the study start date for each of our analyses.

### Statistical methods

#### Kaplan-Meier Survival Curves

For our first analysis, the Kaplan-Meier survival function was estimated from 23^rd^ March to 14^th^ June for each year of care home residency (2016 – 2020).

#### Cox regression

Cox regression was used to determine hazard ratios (HRs) for mortality with 95% confidence intervals. Adjusted hazard ratios included: the cohort year, care home residency, age, gender, HFRS and WIMD (2019 version). We included a cluster level effect for each residence. Computation restrictions meant we were unable to include a cluster level effect for the second analysis.

## RESULTS

### Research question 1: How does mortality in care homes for older adults compare between 2016-2020?

We analysed over 12,000 individuals per year in more than 500 care homes for older adults. We present the descriptive data for the cohorts in Table 1, Kaplan-Meier survival curve in Figure 1 and Cox proportional hazards models in Table 2.

**Table 1.** Demographic information for each of the care home cohorts stratified by year.

| Cohort Year | 2016 | 2017 | 2018 | 2019 | 2020 |
| --- | --- | --- | --- | --- | --- |
| *Individuals (N)* | 13950 | 13481 | 12707 | 12642 | 12568 |
| *Care homes* | 572 | 572 | 572 | 554 | 544 |
| *Deaths (n)* | 1103 | 1076 | 1059 | 941 | 1655 |
| *Deaths (%)* | 7.91% | 7.98% | 8.33% | 7.44% | 13.17% |
| Age |  |  |  |  |  |
| *Mean age (sd)* | 85.3 (8.4) | 85.3 (8.3) | 85.2 (8.4) | 85.2 (8.4) | 85.2 (8.5) |
| Gender |  |  |  |  |  |
| *Female* | 72.0% | 71.5% | 71.4% | 71.5% | 70.8% |
| HFRS |  |  |  |  |  |
| *No score* | 40.0% | 39.8% | 40.2% | 39.9% | 40.8% |
| *Low* | 10.3% | 9.7% | 9.2% | 8.9% | 9.0% |
| *Intermediate* | 25.2% | 24.6% | 24.0% | 23.6% | 23.3% |
| *High* | 24.4% | 25.8% | 26.6% | 27.7% | 26.9% |
| WIMD 2019 |  |  |  |  |  |
| *1. Most deprived* | 15.5% | 15.4% | 15.3% | 15.7% | 15.2% |
| *2.* | 22.0% | 22.0% | 21.3% | 21.8% | 21.8% |
| *3.* | 22.1% | 22.3% | 21.8% | 21.4% | 21.1% |
| *4.* | 20.1% | 20.6% | 20.8% | 20.2% | 20.8% |
| *5. Least deprived* | 20.3% | 19.7% | 20.8% | 21.0% | 21.1% |

**Table 2.** The table shows the Hazard Ratios for adjusted Cox proportional hazards models with a cluster effect for each care home.

| Models (aggregate inclusion) | Cohort year | Age | Gender | HFRS | WIMD |
| --- | --- | --- | --- | --- | --- |
| Cohort year (baseline: 2016) |  |  |  |  |  |
| 2017 | 1.01<br>(0.93,1.09) | 1.01<br>(0.93,1.10) | 1.01<br>(0.93,1.09) | 1.00<br>(0.93,1.09) | 1.00<br>(0.93,1.09) |
| 2018 | 1.06<br>(0.97,1.15) | 1.06<br>(0.97,1.16) | 1.06<br>(0.97,1.16) | 1.05<br>(0.97,1.15) | 1.05<br>(0.97,1.15) |
| 2019 | 0.94<br>(0.86,1.03) | 0.94<br>(0.86,1.03) | 0.94<br>(0.86,1.03) | 0.93<br>(0.85,1.01) | 0.93<br>(0.85,1.01) |
| 2020 | 1.72<br>(1.56,1.91) | 1.73<br>(1.56,1.92) | 1.73<br>(1.56,1.91) | 1.72<br>(1.55,1.90) | 1.72<br>(1.55,1.90) |
| Age (continuous 60+) |  |  |  |  |  |
| Age | - | 1.03<br>(1.03,1.04) | 1.04<br>(1.03,1.04) | 1.04<br>(1.03,1.04) | 1.04<br>(1.03,1.04) |
| Gender (baseline: female) |  |  |  |  |  |
| Male | - | - | 1.44<br>(1.35,1.53) | 1.44<br>(1.35,1.53) | 1.39<br>(1.31,1.47) |
| HFRS (baseline: no score) |  |  |  |  |  |
| Low | - | - | - | 1.08<br>(0.96,1.20) | 1.07<br>(0.96,1.20) |
| Intermediate | - | - | - | 1.30<br>(1.21,1.39) | 1.30<br>(1.21,1.39) |
| High | - | - | - | 1.08<br>(0.96,1.20) | 1.64<br>(1.54,1.75) |
| WIMD 2019 (baseline: 1. Most deprived) |  |  |  |  |  |
|  | - | - | - | - | 0.98<br>(0.95,1.00) |
|  | - | - | - | - | - |
| Concordance | 0.555 (s.e.<br>0.005 ) | 0.595 (s.e.<br>0.005 ) | 0.606 (s.e.<br>0.004 ) | 0.621 (s.e.<br>0.004 ) | 0.622 (s.e.<br>0.004 ) |

**Figure 1.**
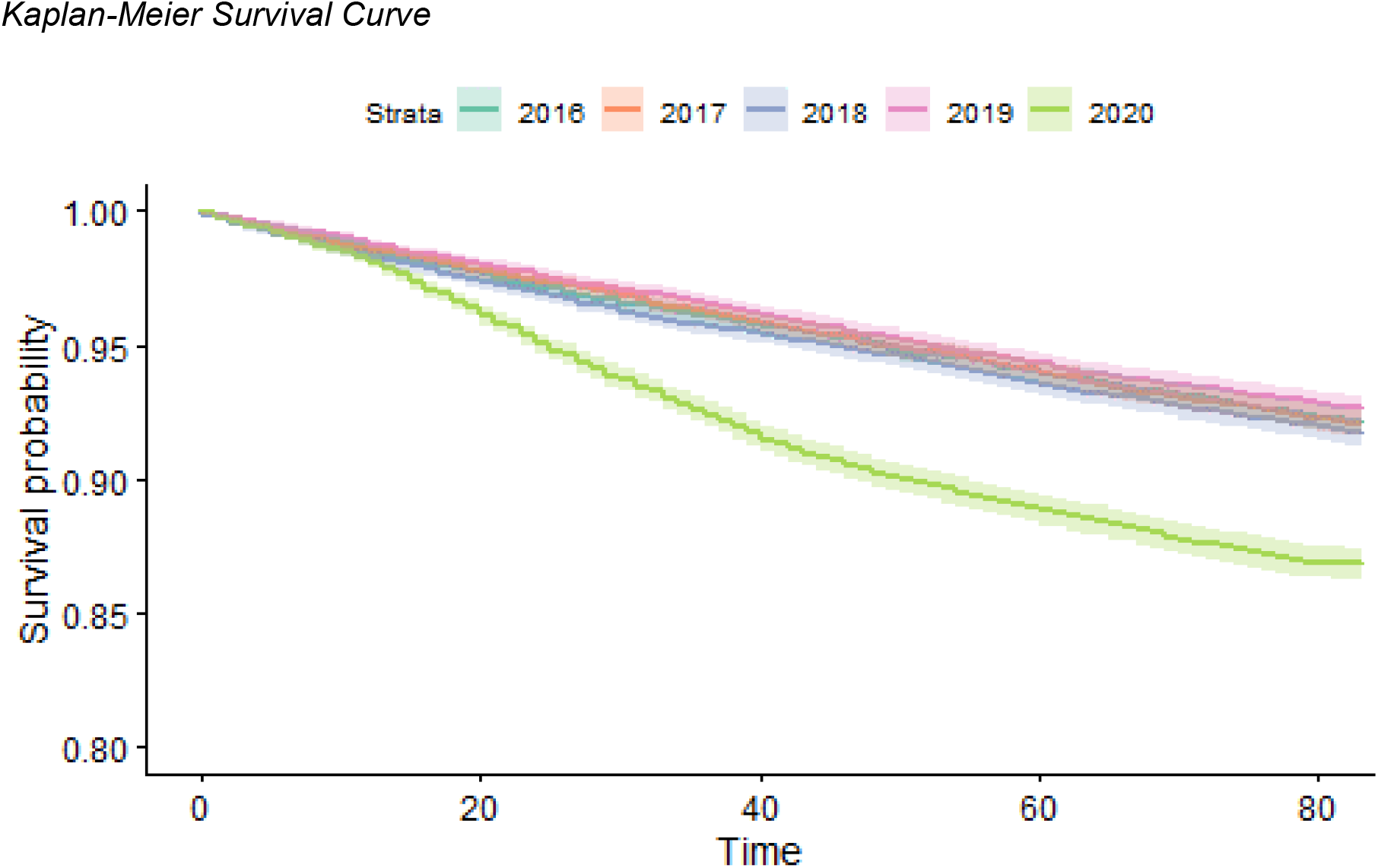
The figure shows the Kaplan-Meier Curves for each cohort (2016-2020). The horizontal axis refers to the time in number of days after the 23^rd^ March up until the 14^th^ June for each year.

### Sensitivity analyses

To check the influence of individuals being present in more than one cohort we have included the number of individuals who are common across each study year in Table S1. We independently calculated adjusted HRs for the 2020 cohort against each of the study years, the results are presented in Table S2. We also present the HRs without a cluster level effect in Table S3.

### Research Question 2: How does care home mortality for older adults compare between 2020 and 2016-2019 in context of the population of Wales?

Our extended analysis included over 3million individuals in the 2016-2019 and 2020 periods of study. The demographic information of each of the cohorts is presented in Table 3 and the corresponding regression model results are displayed in Table 4. Additional models with the individual covariates are presented in Table S4.

**Table 3.**
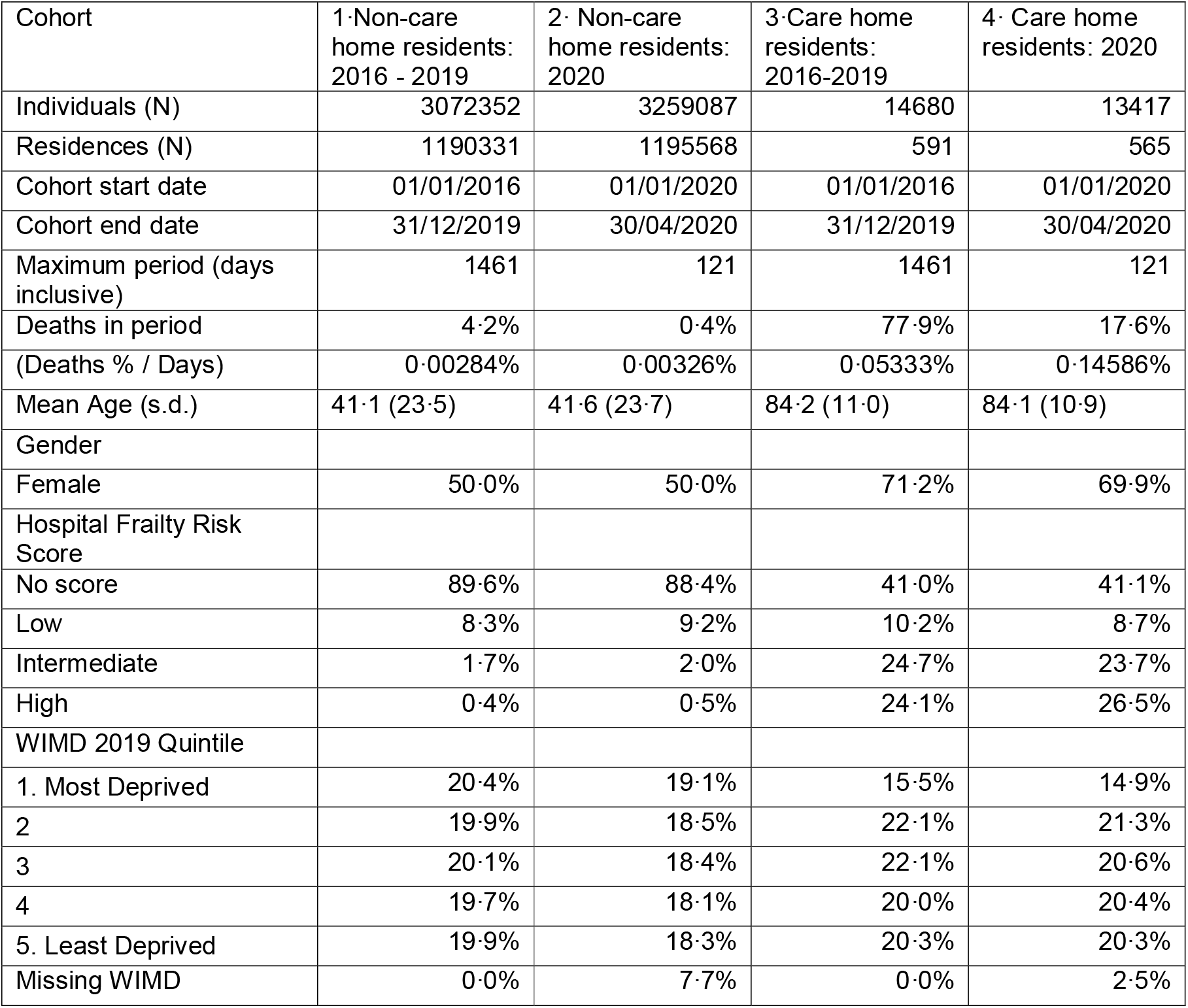
Population wide cohort demographics for care home and non-care home residents in 2020 and 2016-2019.

| Cohort | 1·Non-care home residents: 2016 - 2019 | 2· Non-care home residents: 2020 | 3·Care home residents: 2016-2019 | 4· Care home residents: 2020 |
| --- | --- | --- | --- | --- |
| Individuals (N) | 3072352 | 3259087 | 14680 | 13417 |
| Residences (N) | 1190331 | 1195568 | 591 | 565 |
| Cohort start date | 01/01/2016 | 01/01/2020 | 01/01/2016 | 01/01/2020 |
| Cohort end date | 31/12/2019 | 30/04/2020 | 31/12/2019 | 30/04/2020 |
| Maximum period (days inclusive) | 1461 | 121 | 1461 | 121 |
| Deaths in period | 4·2% | 0·4% | 77·9% | 17·6% |
| (Deaths % / Days) | 0·00284% | 0·00326% | 0·05333% | 0·14586% |
| Mean Age (s.d.) | 41·1 (23·5) | 41·6 (23·7) | 84·2 (11·0) | 84·1 (10·9) |
| Gender |  |  |  |  |
| Female | 50·0% | 50·0% | 71·2% | 69·9% |
| Hospital Frailty Risk Score |  |  |  |  |
| No score | 89·6% | 88·4% | 41·0% | 41·1% |
| Low | 8·3% | 9·2% | 10·2% | 8·7% |
| Intermediate | 1·7% | 2·0% | 24·7% | 23·7% |
| High | 0·4% | 0·5% | 24·1% | 26·5% |
| WIMD 2019 Quintile |  |  |  |  |
| 1. Most Deprived | 20·4% | 19·1% | 15·5% | 14·9% |
| 2 | 19·9% | 18·5% | 22·1% | 21·3% |
| 3 | 20·1% | 18·4% | 22·1% | 20·6% |
| 4 | 19·7% | 18·1% | 20·0% | 20·4% |
| 5. Least Deprived | 19·9% | 18·3% | 20·3% | 20·3% |
| Missing WIMD | 0·0% | 7·7% | 0·0% | 2·5% |

**Table 4.**
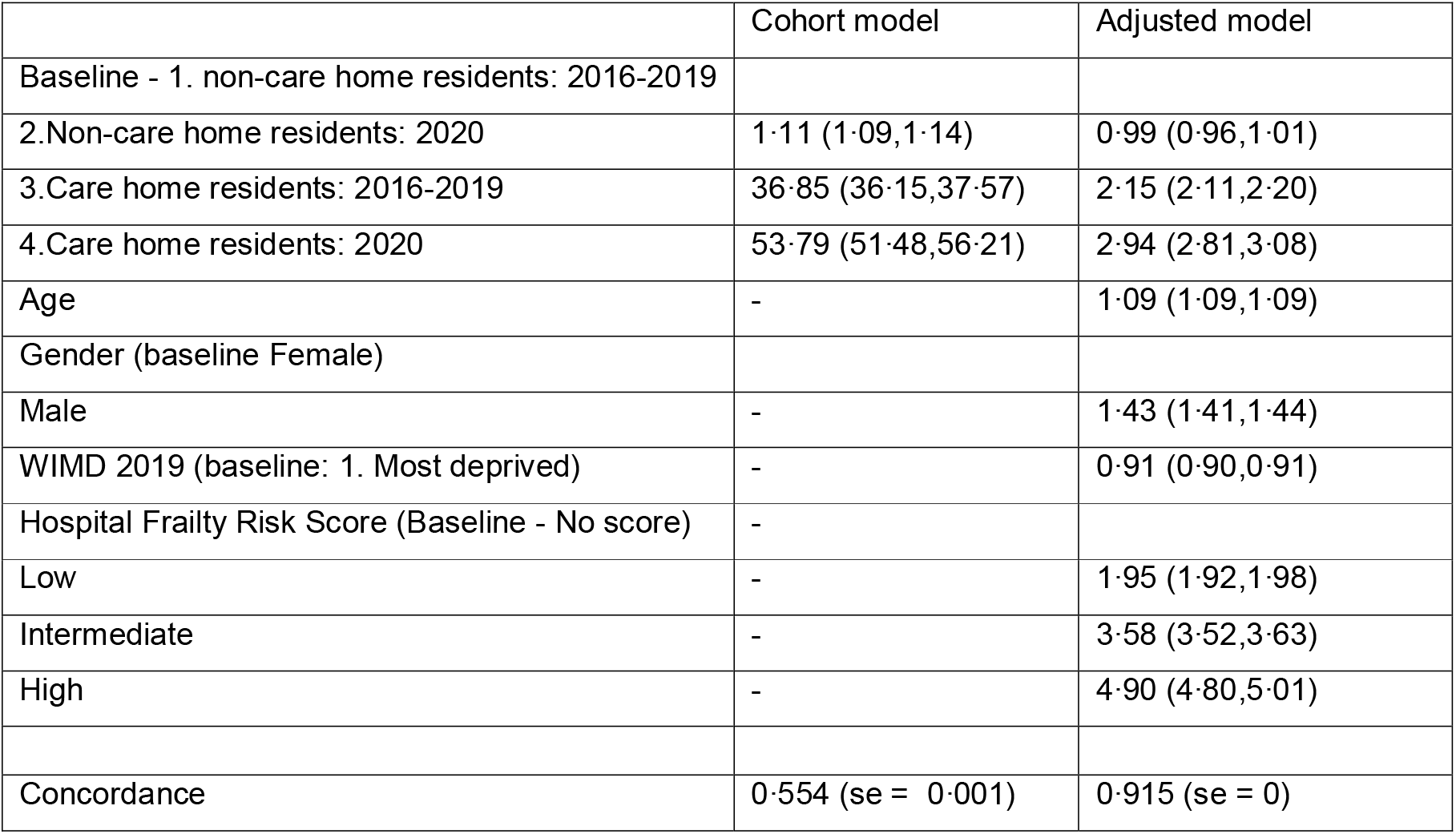
Cox regression results for the extended observation period. Adjustments for age, gender, WIMD and the HFRS are included. The baseline cohort was non-care home residents in the period 2016-2019.

## DISCUSSION

Our results show a substantial excess mortality and substantial reduction in survival in care home residents during the first phase of the lockdown period, when compared to previous years and after adjustment for age, sex, deprivation and hospital frailty risk score. The baseline demographics shown in Table 1 show a consistent trend across each study year with the exception of mortality in 2020. This is consistent with the diverging Kaplan-Meier curves displayed in Figure 1 and the increased hazard ratios for the cohort year presented in Table 2. The HR for the cohort year remains statistically insignificant for 2017/18/19 when compared to 2016, but the HR for 2020 is consistently greater than 1·7 for each of the models presented. We adjusted for age, gender, HFRS and WIMD in the models. It was found that age, gender (male), and increasing HFRS led to an increased HR for mortality. This is consistent with previous studies, where frailty has been shown to be a determinant of mortality [23]–[25]. We found that the WIMD was statistically insignificant and did not alter the remaining HRs for mortality when included in the first analysis, but was a significant factor in the population level analyses. The cluster effect term indicated there was variation between care homes, this is likely because of differences in the case mix of care home residents and the varying exposure to COVID cases.

The KM curves may indicate a flattening of the divergence in mortality in more recent weeks. We plan to repeat these analyses as more data become available. Inclusion of data on the timing of interventions and policy changes, both across the health care system and in care homes would help understand the effectiveness of different approaches on reducing transmission of infection and clinical outcomes.

Table 3 details the differences in demographic information between care home residents and the general population. Specifically, the care home residents are more likely to be women and have increased mortality, frailty and age. The analysis in Table 4 indicated a higher risk of mortality for care home residents compared to the general population. The analysis also showed an increased risk of mortality in care homes in 2020 compared to the 2016-2019 counterparts. Interestingly, the mortality risk of the general population in 2020 compared to 2016-2019 was statistically insignificant, further highlighting the increased risk of mortality in care homes in 2020.

### Limitations

Although we used a consistent list of anonymised care home addresses there is a varying number of care homes included in each year of study. This is due to the list of care homes being created from the 2018 extract from CIW, and care homes being opened and closed. We aimed to mitigate bias in our comparisons by using a consistent list across study years.

Our cohorts were created at cross sectional time points, this means that individuals may appear in more than one cohort. Although we calculated the covariates at the individual level at the start of each cohort interval, there may still remain correlation between the cohorts.

## CONCLUSIONS

We performed a retrospective population-based cohort study, comparing the mortality risk in care homes between 2016 – 2020. It was found that the mortality risk in care homes has increased significantly in 2020 compared to previous years. The conclusion of increased mortality risk in 2020 remained the same when we included additional demographic variables, the HFRS, and increased the observation window.

## Data Availability

The data used in this study are available in the SAIL Databank at Swansea University, Swansea, UK. All proposals to use SAIL data are subject to review by an independent Information Governance Review Panel (IGRP). Before any data can be accessed, approval must be given by the IGRP. The IGRP gives careful consideration to each project to ensure proper and appropriate use of SAIL data. When access has been approved, it is gained through a privacy-protecting safe haven and remote access system referred to as the SAIL Gateway. SAIL has established an application process to be followed by anyone who would like to access data via SAIL https://www.saildatabank.com/application-process.

## Acknowledgements

This work uses data provided by patients and collected by the NHS as part of their care and support. We would also like to acknowledge all data providers who make anonymised data available for research.

We wish to acknowledge the collaborative partnership that enabled acquisition and access to the de-identified data, which led to this output. The collaboration was led by the Swansea University Health Data Research UK team under the direction of the Welsh Government Technical Advisory Cell (TAC) and includes the following groups and organisations: the Secure Anonymised Information Linkage (SAIL) Databank, Administrative Data Research (ADR) Wales, NHS Wales Informatics Service (NWIS), Public Health Wales, NHS Shared Services and the Welsh Ambulance Service Trust (WAST).

## Ethics

This study has been approved by the IGRP as project 0911.

## Funding acknowledgements

This work was supported by Health and Care research Wales [Project: SCF-18-1504]; and Health Data Research UK [NIWA1] which receives its funding from HDR UK Ltd funded by the UK Medical Research Council, Engineering and Physical Sciences Research Council, Economic and Social Research Council, Department of Health and Social Care (England), Chief Scientist Office of the Scottish Government Health and Social Care Directorates, Health and Social Care Research and Development Division (Welsh Government), Public Health Agency (Northern Ireland), British Heart Foundation (BHF) and the Wellcome Trust.

This research has also been supported by the ADR Wales programme of work. The ADR Wales programme of work is aligned to the priority themes as identified in the Welsh Government’s national strategy: Prosperity for All. ADR Wales brings together data science experts at Swansea University Medical School, staff from the Wales Institute of Social and Economic Research, Data and Methods (WISERD) at Cardiff University and specialist teams within the Welsh Government to develop new evidence which supports Prosperity for All by using the SAIL Databank at Swansea University, to link and analyse anonymised data. ADR Wales is part of the Economic and Social Research Council (part of UK Research and Innovation) funded ADR UK (grant ES/S007393/1).

